# Coproducing a conceptual understanding of unmet palliative care needs: stakeholder workshops using modified nominal group technique

**DOI:** 10.1101/2025.08.08.25333284

**Authors:** AE Bone, M Diggle, T Johansson, A Finucane, KE Sleeman, JM Davies, IJ Higginson, LK Fraser, FEM Murtagh

## Abstract

**Background:** Despite growing recognition of the importance of identifying unmet needs in palliative care, there remains no clear, operational definition of what constitutes unmet palliative care needs. This gap hinders service planning, policy development and equitable access to care. We aimed to explore priorities for understanding and measuring unmet palliative care needs with stakeholders, including people with lived and professional experience.

**Methods:** Three online workshops using a modified nominal group technique with people with lived experience of life-limiting illness and professionals in palliative care. Separate workshops were held for each group to independently identify elements that capture the concept of unmet palliative care needs, followed by a combined workshop to refine and consolidate findings. Workshop data were analysed using content analysis. Participants then completed an online ranking exercise to prioritise key elements, which was analysed descriptively.

**Findings:** Twenty-eight individuals participated, including 11 people with lived experience and 17 with professional experience. In the final workshop, participants agreed on a list of 27 elements capturing unmet palliative care needs, which were conceptualised in two main ways: 1) service-related (e.g. lack of a single point of contact to access support, including out-of-hours), and those related to symptoms and concerns, (e.g. pain not assessed and managed). Twenty-three participants completed the ranking exercise. Highly prioritised elements included lack of timely and holistic assessment of symptoms or suffering, inability to access services needed, lack of coordination, and continuity of care. Other priorities for understanding and measuring unmet palliative care needs were lack of timely follow-up to address symptoms, lack of skilled support, and lack of respect, dignity and empathy.

**Conclusions:** This study is the first to engage both individuals with lived experience and professionals in conceptualising unmet palliative care needs. Stakeholders prioritised two broad aspects important for capturing unmet palliative care needs: symptom-related concerns and service-related issues. These findings provide a foundation for developing stakeholder-informed tools to assess unmet palliative care needs that are feasible for use across diverse care settings and populations.

## Background

The need for palliative care is a growing issue globally due to ageing populations and rising serious health related suffering.(1) *Unmet* palliative care needs can be conceptualised as the gap between required or expected care and actual care received by people with life-limiting illness.(2) Unmet palliative care needs are a concern at multiple levels: for individuals and their families who are experiencing unresolved symptoms and concerns; for healthcare professionals and service managers to understand which issues to target; for policymakers and commissioners who plan and commission services to meet population needs; and for society, due to the adverse societal impact of unmet needs, such as burden and costs associated with informal care and prolonged or complicated grief.(3)

Estimating unmet palliative care needs is crucial for identifying where services fall short in supporting this vulnerable population, and for determining necessary service improvements. However, quantifying unmet palliative care needs is challenging, primarily due to the lack of an accepted operational definition.(4) This lack of definitional clarity hampers efforts to compare findings across studies, evaluate service effectiveness, and design interventions that address unmet needs. Our scoping review of the literature revealed that despite being widely studied, unmet palliative care needs were generally poorly defined and inconsistently measured and reported.(2) Although person-centered care is a cornerstone of modern healthcare,(5) our scoping review found little evidence that stakeholders - particularly those with lived experience - have been consulted in defining or measuring unmet palliative care needs. Engaging stakeholders with lived and professional experience is essential to ensure that any operational definition reflects the real-world complexity of palliative care needs and aligns with the principles of person-centered care.

In this study we aimed to explore priorities for understanding and measuring unmet palliative care needs with stakeholders, including people with i) lived experience of life-limiting illness (people with life-limiting illness and their informal family caregivers), and ii) professional experience (including delivery, planning, and commissioning care for people with life-limiting illness), to inform an operational definition of the construct.

## Methods

### Design

This study is part of a wider project titled *Defining and Estimating Unmet Palliative Care need: DUECare project* (grant MCCRP-23-02). We report here a series of workshops with people with lived and professional experience, guided by a modified nominal group technique(6, 7). This structured method is useful for generating ideas and establishing areas of agreement among a diverse group of individuals in a short period of time.(8, 9) It is also a highly adaptable method to suit the needs of the research aim and the participants.(10) We adapted this established method for our aim, by:

1. holding separate workshops specific for the two groups: i) those with lived experience of life-limiting illness, and ii) those with professional experience. This was to ensure both groups had the opportunity to independently generate ideas on unmet palliative care needs, without influence from the other group.
2. conducting a combined workshop with individuals involved in the first two workshops to work towards consensus, including a subsequent ranking exercise, to establish the priorities of individual participants for understanding and measuring unmet palliative care needs.

The qualitative aspects of the study were reported according to the Consolidated Criteria for Reporting Qualitative research (COREQ) guidelines (Appendix Table 1).(11)

### Setting and Participants

We included the following groups from across the United Kingdom:

1. people aged 18 or older with lived experience of having life-limiting illness or caring for an adult with life-limiting illness and;
2. people with professional experience of delivering, planning or commissioning health and social care for people with life-limiting illness.

People with lived experience were identified using established patient and public involvement (PPI) networks from a range of locations to maximise diversity of views, including the Cicely Saunders Institute online PPI forum; the PPI network INVOLVE Hull; and the Marie Curie Voices network. We posted an advertisement through these networks requesting that interested individuals contact researchers by email or telephone to arrange a time to talk about the workshops in more detail. The participant information sheet, outlining the study aim and potential risks and details about participation and data management, was shared with those who were interested.

We purposively sampled people with professional experience of delivering, planning or commissioning services for people with life-limiting illness, to gain a diversity of views across different roles (doctors, nurses, allied healthcare, service managers, commissioners). We identified relevant individuals through the researchers’ professional networks and contacted them via email, including the participant information sheet. Where professionals were interested but unable to attend, they were asked to nominate colleagues with similar experience. When a potential participant expressed interest in participating, one of the research team offered to discuss the study with them. This step ensured potential participants fully understood the purpose of their participation and could ask any questions. On receipt of a signed consent form, participants were asked to complete a brief demographic survey so the characteristics of the sample could be described.

### Data collection

We held three online workshops via Microsoft Teams in January and February 2025, each lasting three hours including breaks. Table 1 provides details of the procedures following modified Nominal Group Technique.(6, 7) The first two workshops explored what the stakeholders (people with lived experience, and people with professional experience) considered to be the key aspects of unmet palliative care needs, each following the same format. During these workshops, scribes made notes from each group discussion and the wider discussions which were used to derive elements of unmet palliative care needs. All workshops were also recorded to facilitate accurate note checking. The third workshop included both participant groups. In this workshop, the research team presented the initial list of elements of unmet palliative care needs for discussion, elaboration and refinement. This included combining overlapping areas, adding new elements, and amending language for clarification. Stakeholders agreed on the final list of elements of unmet palliative care needs (without prioritising or ranking at this stage). Participants were encouraged to email with any further thoughts and reflections which were captured as research notes.

**Table 1.** Steps of the modified nominal group technique.

| Activity | Mode* | Date |
| --- | --- | --- |
| 1. Brief presentation of findings from a scoping review on defining and measuring unmet palliative care needs. | Workshops 1 and 2 | Jan 2025 |
| 2. In small, facilitated groups participants discussed the following questions:<br>- *What does good care look like for people with life-limiting illness?*<br>- *What are the most important gaps between the care needed and care that is received?*<br>- *Are some symptoms or problems more important for understanding the gaps in care than others?*<br>- *Can we understand whether needs have been met from the services someone accessed?*<br>- *What is the best time period for understanding the gaps in care?* | Workshops 1 and 2 |  |
| 3. Key discussion points were fed back to the whole group for further discussion, clarification and consolidation. | Workshops 1 and 2 |  |
| 4. List of elements of unmet palliative care need generated by the research team based on the findings from workshops 1 and 2 | After workshops 1 and 2 | Jan/Feb 2025 |
| 5. Participants were presented with the inductively derived elements of unmet palliative care needs | Workshop 3 | Feb 2025 |
| 6. In small, facilitated groups participants were asked to reflect on the elements of unmet palliative care needs, first in silence. | Workshop 3 |  |
| 7. Each participant was then asked in turn to suggest any necessary amendments to wording or additions to the list if important areas were missed. | Workshop 3 |  |
| 8. Key points were fed back to the wider group and duplicates were identified and overlapping areas were combined. | Workshop 3 |  |
| 9. Group agreement on the final list of elements of unmet palliative care needs. | Workshop 3 |  |
| 10. Ranking exercise to identify the **top 10** most important elements. | Qualtrics software sent via email after workshop 3 | March 2025 |
| **Bold** indicate where the study procedures were modified during the project in response to participant feedback.<br>*Workshop 1 included people with lived experience of life-limiting illness or caring for an adult with life-limiting illness, workshop 2 included people with professional experience of delivering, planning or commissioning health and social care for people with life-limiting illness, and workshop 3 combined participants from workshops 1 and 2. |  |  |

**Table 1.** Workshop participants’ (n=28) characteristics.

|  | People with lived experience N=11 |  | Health and social care professional N=17 |  |
| --- | --- | --- | --- | --- |
|  | Workshop 1 (n=11) | Workshop 3 (n=8) | Workshop 2 (n=16) | Workshop 3 (n=13*) |
| **Gender** |  |  |  |  |
| Male | 4 | 4 | 3 | 2 |
| Female | 7 | 4 | 13 | 11 |
| **Age** |  |  |  |  |
| 18-44 years | 1 | 0 | 3 | 2 |
| 45-64 years | 6 | 4 | 13 | 11 |
| 65 years or over | 4 | 4 | 0 | 0 |
| **Country of residence** |  |  |  |  |
| England | 10 | 7 | 7 | 7 |
| Wales | 0 | 0 | 2 | 1 |
| Scotland | 1 | 1 | 4 | 4 |
| Northern Ireland | 0 | 0 | 3 | 1 |
| **Ethnic group** |  |  |  |  |
| Mixed/Multiple ethnic groups | 3 | 1 | 0 | 0 |
| Asian/Asian British | 2 | 2 | 0 | 1 |
| Black/ African/ Caribbean/ Black British | 0 | 0 | 1 | 1 |
| White | 6 | 5 | 15 | 11 |
| **Experience** |  |  |  |  |
| Informal caregiver | 9 | 6 | N/A | N/A |
| Patient | 2 | 2 | N/A | N/A |
| **Professional role/area** |  |  |  |  |
| Specialist palliative care (physician or nurse) | N/A | N/A | 5 | 3 |
| General practice | N/A | N/A | 1 | 1 |
| Allied health | N/A | N/A | 2 | 2 |
| Pharmacy | N/A | N/A | 1 | 1 |
| Public health | N/A | N/A | 1 | 1 |
| Policy/strategy | N/A | N/A | 4 | 3 |
| Social care | N/A | N/A | 1 | 1 |
| Commissioner | N/A | N/A | 1 | 1 |
| Mean Years in role | N/A | N/A | 7.7 years | 8.4 years |
| *\*Includes one professional who was unable to attend workshop 2* |  |  |  |  |

Finally, as part of the modified nominal group technique, participants were invited to prioritise elements that can capture unmet palliative care needs through a ranking exercise using *Qualtrics* software (Qualtrics, Provo, UT). The protocol detailed that participants would be asked to rank their top three most important elements to work towards consensus and inform measurement of unmet palliative care needs.

In response to participant feedback during the project, the research team made modifications to the research procedures to maximise meaningful engagement. Firstly, some participants reported that prioritising their top three elements of unmet palliative care need was too difficult, as they felt this would not reflect the breadth of issues. Secondly, participants raised concerns about the ranking exercise during the workshop without sufficient time for reflection. In response, the study team postponed the ranking exercise to revise the instructions and allow more time for reflection. With the participants’ agreement and ethics committee approval, an online version of the ranking exercise was later sent to participants. We asked participants to rank the top ten elements instead of the top three.

Participants were encouraged to include free text comments to provide further details on their prioritisation.

### Analysis

Discussions from workshops 1 and 2 were captured as notes and analysed using content analysis at a semantic level to inductively derive elements of unmet palliative care needs.(12) We selected content analysis as a useful approach to determine the words and phrases which identified the various domains, elements, or concepts relating to areas of unmet palliative care need; it is largely descriptive and has the advantage of remaining close the ideas communicated by participants.(13)

The analysis drew on the workshop discussions responding to questions *What does good care look like for people with life-limiting illness?* and *What are the most important gaps between the care needed and care that is received?* We adopted a pragmatic approach to allow for rapid synthesis of findings from workshops 1 and 2, informing the third workshop within a short timeframe. In studies using nominal group technique, this synthesis is usually carried out within stakeholder workshops, however, this additional step allowed participants more time to reflect on the elements identified in order to rank them.(10) The inductively derived elements were informed by the frequency and depth of discussion across the two workshops, with divergent views also captured. The analysis was discussed within the research team to ensure they reflected the workshop discussions and captured nuances.

The research team invited those participants who had contributed to the workshops to take part in the online ranking exercise. Ranks were assigned numbers to weight for importance (e.g. rank 1 = 10 points, rank 2 = 9 points etc).(14) We analysed the scores in Excel by summing the individual rank scores.(10) For each item we also presented the proportion included in the respondents’ top 10.(15)

### Ethical considerations

Ethical approval was sought from King’s College London and received full approval on 6^th^ December 2024 (LRS/DP-24/25-45413). Written informed consent was gained from all participants via email before the start of the workshops. People with lived experience were reimbursed for their time in line with National Institute for Health and Care Research (NIHR) public contributor payment policy.(16)

## Findings

In total 28 individuals participated in the study (table 1). The research team received initial responses from 20 people with lived experience, of whom 11 participated in the first workshop. Six did not respond to subsequent emails, two did not meet the eligibility criteria and one declined. The research team approached 30 health and social care professionals; 17 expressed an interest in taking part while four declined and nine did not respond. 16 health and social care professionals took part in workshop 2; one declined workshop 2 due to availability but participated in workshop 3. Of the 28 individuals who joined workshops 1 and 2, 21 participated in the third workshop, including eight people with lived experience and 13 health and social care professionals (Table 1).

### Workshops 1 and 2

The discussions in workshops 1 and 2 encompassed how people with lived and professional experience conceptualised unmet palliative care needs, and which domains, elements and concepts they used which reflected this. Findings from these workshops are presented in relation to the open questions participants were asked to consider.

#### What does good care look like for people with a life-limiting illness?

In relation to good care, participants emphasised relational aspects of care, identifying themes such as person-centredness, being treated with empathy and respect. Participants described the importance of being seen as an individual and having a responsive service that listens and adapts to what is important to that person. Another prominent theme was the important role health and social care professionals can play in supporting dignity in care.

Participants considered that skilled and competent care from professionals was crucial, including early assessment of problems and concerns, and timely action, particularly in reference to pain relief. This was needed in order to foster trust between patients/families and professionals. Participants also highlighted the importance of knowing who to contact and how (for example accessible advice lines), particularly out of hours.

#### What are the important gaps between care needed and care received?

Participants emphasised that a critical gap in care for people with life-limiting illness is the lack of recognition of a person’s nearness to end of life. It was suggested that deficits in active listening in clinical care may result in palliative care needs going unrecognised. Access to care and medications out of hours was also emphasised as a big gap in care leading to unmet palliative care needs, and timely access to equipment in the home. Challenges in navigating the care system when there is no single point of contact was considered to be another important gap. Without continuity of care participants expressed concerns about the ability of health and social care professionals to provide personalised care and foster trust from individuals and their families. Also highlighted across the groups was the importance of competence and skilled health care professionals to avoid failures of care, which may lead to partially or poorly met palliative care needs.

It was noted that it is important to consider outcomes (e.g. unresolved symptoms) and not just processes (e.g. lack of access to services). Participants highlighted that unresolved pain is an important unmet palliative care need. However, it was also argued that pain is often the focus of care providers at the expense of other symptoms, such as anxiety and depression which do not receive enough attention. Financial concerns and practical matters such as legal issues and information were also highlighted as important areas of unmet palliative care need.

#### Are some symptoms or problems more important for understanding the gaps in care than others?

Many participants agreed that gaps in care can only be determined by the individual and what matters to them and that this can change over time. Nevertheless, a hierarchy of symptoms was apparent, with pain, anxiety, breathlessness, and nausea being consistently highlighted as most significant. There was also the suggestion that as a person nears the end of life, the psychological, spiritual and social needs become more significant but are less well met than physical needs. Participants across both workshops emphasised the importance of “basic” needs, which are required to maintain dignity. These include physical comfort afforded by adequate housing, nutrition, heating, and ‘basic care’ including personal care. Participants suggested that the theoretical framework of Maslow’s hierarchy of needs(17) clarifies the importance of different needs and their interconnectedness, with basic needs being foundational before higher-level needs can be met. Moreover, it was noted that presence of symptoms or problems does not necessarily reflect whether a service is wanted or appropriate for the individual.

#### Can we understand whether needs have been met from the services someone accessed?

Most agreed that simply receiving a service does not necessarily imply that needs are met. Conversely, not receiving specialist palliative care does not equate to unmet needs. Although some suggested that certain patterns of service use such as high frequency visits to the emergency department, could indicate that needs were not being adequately met, this was recognised as an imperfect indicator since urgent care service use may be appropriate. One participant posited that understanding unmet palliative care needs does not require information about access to services or symptoms, rather it is establishing whether an individual receives care that they perceive is important and appropriate to them.

The workshop discussions also addressed which services and professionals should be addressing unmet palliative care needs. Some expressed that everyone should receive generalist palliative care provided by their usual health and social care teams, and access to specialist palliative care should be available for those with complex care needs. Others pointed out the inequities in service provision, with the ‘privileged few’ receiving specialist palliative care services, whereas other more marginalised groups have less access to both specialist and generalist services, such as those with severe mental illness, those experiencing homelessness, and those aged over 85-years with multiple conditions who are more likely to experience unmet needs.

#### What is the best time period for understanding the gaps in care?

Participants emphasised that palliative care needs are dynamic, changing over time, and are unique to the person. Determining the optimal time period to collect information about unmet palliative care needs at a population level was therefore considered difficult. The last year of life was deemed a preferable timeframe but potentially impractical. Several participants agreed that the last three months of life is usually when care is most intensely needed and might be an appropriate time period for understanding unmet palliative care needs.

It was noted that ascertaining unmet palliative care needs from patient and carer reported information can be challenging. For example, some individuals have difficulty in expressing their needs for a variety of reasons, including language issues, impaired cognition, lack of awareness, an unwillingness to acknowledge difficulties, perceived stigma, or concerns about upsetting family members. Participants also pointed out that unmet needs might not be recognised if patients and families have limited awareness of which services are available, what they are entitled to, or what services can address.

From the workshop discussions the research team derived an initial list of 21 important elements of unmet palliative care needs (Appendix table 2). There were two broad ways of conceptualising unmet palliative care need – unaddressed symptoms and concerns (e.g. *unaddressed pain*; *unaddressed breathlessness*; and *unaddressed depression and anxiety, and unaddressed spiritual needs)*, and lack of timely and high-quality services received (e.g. *lack of timely assessment of what is important to the person; inability to access services needed; lack of coordination and continuity of care)*.

### Workshop 3

The 21 elements capturing unmet palliative care needs were presented at the third workshop and revised with input from stakeholders. Language was amended to increase clarity. For example, *Unaddressed pain* was expanded to *Pain not assessed and managed.* In some cases, elements were combined, for example *Unaddressed depression* and *unaddressed anxiety* became *Depression or anxiety not assessed and managed.* Seven new elements were added, including *Lack of access to skilled and competent health and social care professionals* and *Unaddressed distress (e.g. existential/ fear of dying).* Participants agreed on the final list of 27 elements of unmet palliative care needs.

### Ranking of elements of unmet palliative care needs

Twenty-six participants who had contributed to the workshops were invited to the online ranking exercise. All 26 responded, with 23 participants completing the ranking and the remaining three opting to provide free text comments only. The aggregated ranking scores are presented in Table 2. *Lack of timely and holistic assessment of symptoms or suffering* received the highest total score, and *Inability to access services needed* was the item most frequently appearing in the top 10 (74%).

**Table 2.** Total rank scores for elements of unmet palliative care needs (N=23).

| Elements of unmet palliative care needs | Total Score | Number (%) in top 10 rank |
| --- | --- | --- |
| Lack of timely and holistic assessment of symptoms or suffering | 92 | 15 (65) |
| Inability to access services needed | 91 | 17 (74) |
| Lack of coordination and continuity of care | 88 | 14 (61) |
| Not being recognised/identified as having palliative care needs | 86 | 14 (61) |
| Lack of a single point of contact to access support, including out-of-hours | 85 | 14 (61) |
| Lack of timely assessment of what is important to the person | 84 | 14 (61) |
| Lack of access to skilled and competent health and social care professionals | 76 | 11 (48) |
| Not being treated with respect, dignity and empathy | 64 | 11 (48) |
| Lack of satisfactory timely follow-up action after assessment | 59 | 8 (35) |
| No discussion or planning for future care | 52 | 12 (52) |
| Lack of timely access to medicines | 51 | 9 (39) |
| Pain not assessed and managed | 50 | 8 (35) |
| Lack of referral by self or professionals to specialist palliative care if needed | 50 | 11 (48) |
| Unaddressed housing, financial, legal or other practical issues | 44 | 10 (43) |
| Lack of understandable information about the disease & treatment choices | 43 | 9 (39) |
| Lack of understandable information about services and support available | 40 | 8 (35) |
| Lack of help with basic care (e.g. personal care) | 37 | 7 (30) |
| Being unsafe | 32 | 4 (17) |
| Unaddressed existential fear or distress | 32 | 5 (22) |
| Carers/family members unsupported | 30 | 9 (39) |
| Breathlessness not assessed and managed | 18 | 2 (9) |
| Lacking support and equipment for functional independence | 18 | 6 (26) |
| Other physical symptoms not assessed and managed | 15 | 4 (17) |
| Unaddressed cultural or ritual needs | 15 | 4 (17) |
| Depression or anxiety not assessed and managed | 7 | 2 (9) |
| Unaddressed spiritual or religious needs | 6 | 2 (9) |

In total, 16 respondents provided free text comments to describe their reasoning for prioritisation. Some noted that certain elements were broad and could be considered to encompass others and which led some to rank these elements highly to capture more aspects of unmet needs that they felt were important. For example, one participant wrote they had “*not included any specific symptoms as the question about holistic assessment followed by a plan that is actioned would encompass these”*. Another noted that they ranked elements with an equity lens, choosing to prioritise timely identification of palliative care needs at the top because if this is not done equitably, then it further entrenches inequities further downstream.

A few participants noted challenges with completing the ranking exercise. Some felt that the instruction to rank according to importance lacked clarity, for example importance to whom - themselves, professionals, or the public? Another participant was opposed to the concept of ranking elements which to them was *“counter to the idea of total pain*”. Others noted that the elements were interrelated and given they were not mutually exclusive it was difficult to assess them against each other. One participant considered all elements equally important, while others pointed to the fact that the importance of elements will vary by individuals according to their specific situation.

## Discussion

To our knowledge, this is the first study to engage both individuals with lived experience and professionals in exploring how unmet palliative care needs can be defined and measured.(2) Our earlier scoping review revealed that, despite a substantial body of literature, the construct of unmet palliative care needs remains poorly defined, with limited involvement of stakeholders in shaping its conceptualisation and operationalisation.(2) This study advances the field by offering insights into how stakeholders conceptualise unmet palliative care needs and what they view as priorities for measurement.

The most highly prioritised elements capturing unmet palliative care needs focused on service delivery, quality and access, and tended to be broader in scope for example: lack of timely and holistic assessment of symptoms or suffering, inability to access services needed and lack of coordination and continuity of care. Other key priorities identified were the lack of recognition of palliative care needs, absence of a single point of contact for support (including out-of-hours care), lack of timely follow up to address symptoms and concerns, insufficient skilled and competent care, and not being treated with respect, dignity, and empathy. These findings highlight critical areas that should be captured when measuring unmet palliative care needs in care practice and research, and provide a foundation for developing more responsive and stakeholder-informed assessment tools.

Stakeholders identified a wide range of symptoms, concerns, and aspects of service delivery including access and quality of care as areas that reflect unmet palliative care needs, underscoring the complexity of the construct.(18) The breadth of domains is consistent with previous stakeholder engagement on related topics.(19, 20) For instance, studies involving individuals with lived experience and professionals have highlighted social and psychological support, financial concerns, and issues related to coordination, communication, and continuity of care as key to the assessment of palliative care needs.(19, 21)

The elements identified in our study also overlap with domains captured in existing tools such as the Supportive Care Needs Survey - Short Form (SCNS-SF34)(22, 23), the Problems and Needs in Palliative Care – short version (PNPC-sv)(24), and Integrated Palliative Outcome Scale (IPOS)(25, 26), particularly around symptoms, communication, and emotional concerns. However, our findings extend beyond these by highlighting service-level factors—such as access, coordination, and continuity of care—that are less consistently included. Both patient-level factors (such as symptoms and concerns) and service-level factors (such as access, coordination, and continuity) were elements which were highly prioritised by stakeholders and the best measure will depend on the purpose.

Our recent scoping review of the literature on defining and measuring unmet palliative care needs identified three broad approaches to measurement: 1) symptoms and concerns; 2) access to services; and 3) sufficiency of service provision to resolve symptoms and concerns(2). The most highly ranked elements from this study relate to service aspects, which map most closely to access to services and to some extent the sufficiency of service provision. From the ranking exercise, many stakeholders placed more importance on overarching service delivery and quality, such as *lack of timely and holistic assessment of symptoms or suffering* than individual symptoms. The rationale was that if there is timely and holistic assessment of symptoms, this in turn would address a broad range of ‘felt needs’(needs that individuals perceive and express themselves)(27) among people with life-limiting illness. This finding was at odds with some comments in the workshops that outcomes (such as symptom burden) were more important than processes (such as discussing or planning future care). The higher ranking of service aspects rather than individual symptoms may also stem from the fact that the list of symptoms and concerns was not exhaustive. Additionally, prioritising one symptom or domain over another may conflict with the holistic nature of the concept of ‘total pain’(28).

Participants emphasised that unmet palliative care needs relate not only to what care was received and when, but also *how* the care was delivered. Care provided by competent and skilled professionals was perceived as central to fostering trust and a sense of safety - mechanisms previously identified as important for effective palliative care services.(29) Being treated with empathy and respect was also considered crucial in maintaining dignity in the context of life-limiting illness. While the subjective nature of these aspects of care make them more difficult to measure and quantify, our findings emphasise that relational aspects of care are important to capture to better understand unmet palliative care needs.

Stakeholders expressed differing views on which elements of unmet palliative care needs are most important, with only seven elements in over 50% of respondents’ top 10. A strong theme from the workshops was that the relative importance of elements of unmet needs is dependent on what matters to the individual, highlighting that a one-size-fits-all approach to measurement may be inadequate. Some participants supported the idea that evaluating the perceived sufficiency of care, either by the individual or someone close to them, could offer a more person-centred way to address this challenge. This approach has been taken in the development of the Problems and Needs in Palliative Care – short version (PNPC-sv).(24)

### Strengths and Limitations

A key strength of this study was the use of an online workshop format, which removed geographical barriers and enabled participation from individuals with life-limiting illness, as well as clinicians with demanding schedules.(30) Several participants with lived experience also brought prior professional experience in healthcare, while some professionals drew on their own personal experiences of serious illness or caregiving. This overlap enriched the discussions, contributing to a nuanced and multifaceted understanding of the topic.

We purposively sampled professionals to ensure a diverse range of roles and perspectives. However, participants with lived experience were self-selecting and represented a relatively small sample. Our sample included limited representation from some ethnic minority groups, and the extent of diversity across other key characteristics such as socioeconomic position, disability or sexuality remains unclear. Additionally, combining participants with lived and professional experience in the third workshop may have inhibited open discussion for some individuals. Due to the scope and timeframe of the study, there was limited opportunity to explore this complex topic in depth. Our approach to analysis was rapid and pragmatic, which, while appropriate for the study’s aims, may have constrained the depth of interpretation. We aggregated findings across all our participants and did not consider how the importance of different domains of unmet palliative care needs varies across different groups or communities. For example, some minority communities might prioritise rituals and spiritual aspects more highly.(31, 32)

A key methodological challenge for this project in identifying individual elements of unmet palliative care needs for the ranking exercise was defining the scope of each element. Although the research team aimed to create discrete categories, some overlap was inevitable due to the inherently interconnected nature of the issues involved. To better understand the relative value of different measurement approaches, the team combined process and outcome elements within a single ranking exercise. However, this may have introduced cognitive challenges for participants, who were required to evaluate fundamentally different types of information side by side.

### Conclusions

This study provides novel insights into how stakeholders with both lived and professional experience conceptualise unmet palliative care needs. Stakeholders prioritised two broad aspects as important for capturing unmet palliative care needs: symptom-related concerns (e.g. lack of timely and holistic assessment, and unaddressed physical and psychological symptoms) and service-related issues (e.g. lack of timely access, coordination, and continuity of care). Relational aspects of care, such as being treated with dignity and empathy, were also emphasised. Addressing unmet palliative care needs is a shared responsibility across the entire health and social care system, encompassing both generalist and specialist services in various care settings. These findings provide a foundation for developing stakeholder-informed tools to assess unmet palliative care needs that are feasible for use across diverse care settings and populations.

## List of abbreviations

PPI - patient and public involvement

## Declarations

### Ethics approval and consent to participate

Ethical approval was sought from King’s College London and received full approval on 6^th^ December 2024 (LRS/DP-24/25-45413). Written informed consent was gained from all participants via email before the start of the workshops.

### Consent for publication

Not applicable

### Availability of data and materials

The datasets used and/or analysed during the current study are available from the corresponding author on reasonable request.

## Authors’ contributions

AB, KS, IJH, FM, LF, JD, AF contributed to the design and conception of this study. AB, AF, MD, TJ, KS, JD, LF, FM prepared and facilitated the workshops. MD recruited and consented participants to the workshops. AB analysed the data with input from all other authors. AB, AF, MD, TJ, KS, JD, LF, IJH, and FM critically reviewed the manuscript. All authors participated in the process and approved the final version of this manuscript.

## Data Availability

All data produced in the present study are available upon reasonable request to the authors.

## Acknowledgments

We would like to thank all the participants who generously gave their time and shared their experiences for this study.

