## Appendix Table 1 for "Coproducing a conceptual understanding of unmet palliative care needs: stakeholder workshops using modified nominal group technique"

**Appendix Table 1 Consolidated Criteria for Reporting Qualitative Studies (COREQ) guidelines**

| | |
|---|---|
| **Domain 1: Research team and reflexivity** | |
| 1. Interviewer | AEB, KS, AF, and JD facilitated workshop discussions, and FEMM TJ, AF, JD, LF scribed. |
| 2. Credentials | Post doctoral researcher (TJ); Research Fellow (JD); Lecturer (AEB); Senior Research Fellow (AF); Professor (KS, FEMM, LF, IJH). |
| 3. Occupation | Researchers/academics in palliative care, part time clinicians |
| 4. Gender | All females. |
| 5. Experience and training | All members of the research team are postdoctoral researchers. FEM, KS, LF and IJH are professors with substantial research experience. All authors have academic credentials. |
| 6. Relationship with participants | A relationship has been established via email with all participants prior to conducting the workshops. There was a mix of those who were unknown to the research team and those who have relationships with members of the research team through research or clinical activities. |
| 7. Participants knowledge | The research team introduced themselves at the beginning of the workshops and participants were informed about the purpose of the study in a participant information sheet. |
| 8. Interviewer characteristics | Assumptions about unmet palliative care needs were detailed. |
| **Domain 2: study design** | |
| 9. Methodological orientation and Theory | This study used a modified nominal group technique. Documentation from note-takers were analysed using content analysis. |
| 10. Sampling | We used purposive sampling. Where professional experts were interested but unable to attend, they were asked to nominate a colleague to invite. |
| 11. Method of approach | People with lived experience were invited via an advertisement posted on PPI forums and asked to email the research team if they were interested. People with professional experience were approached via email. |
| 12. Sample size | 28 participants |
| 13. Non-participation | People with lived experience: 20 responded to the advert registering interest. Of those, 6 did not respond to subsequent emails, two did not meet the eligibility criteria and one declined. People with professional experience: 30 were invited via email. Of those, 13 declined or did not respond; 1 declined workshop 2 due to availability but participated in workshop 3. |
| 14. Setting of data collection | The workshops were conducted online via Microsoft Teams |
| 15. Presence of non-participants | We had administrative help from one member of the research team who was present to support participants in joining the Microsoft Teams call and be attentive to the well-being of participants. |

| | |
|---|---|
| 16. Description of sample | Participants were described in terms of their self-reported age, gender, ethnicity, country of residence, experience and years of professional experience (where relevant). |
| 17. Interview guide | The workshops followed the steps detailed in Table 1 in the manuscript |
| 18. Repeat interviews | The workshops followed the steps detailed in Table 1 in the manuscript |
| 19. Audio/visual recording | Audio recordings of the workshops were used for noted taking and checking for accuracy only. |
| 20. Field notes | Scribes were assigned to each breakout group to document the discussions |
| 21. Duration | Each workshop was 3 hours in duration. |
| 22. Data saturation | Participants were allowed time to discuss their points in detail and were encouraged to email with any further thoughts after the workshop if there was insufficient time to include them in the workshop. |
| 23. Transcripts returned | Notes from scribes were collated after each workshop for analysis. |
| **Domain 3: analysis and findings** | |
| 24. Number of data coders | Not applicable. |
| 25. Description of the coding tree | Not applicable. |
| 26. Derivation of themes | Themes were inductively derived from the data on the workshop discussions. |
| 27. Software | Not applicable |
| 28. Participant checking | Participants fed back on the elements of unmet palliative care need as per the method detailed in Table 1. |
| 29. Quotations presented | Quotations are not presented. |
| 30. Data findings consistent | Yes, the findings were consistent with the data. |
| 31. Clarity of major themes | Major themes are presented in the findings and repeated in the conclusion. |
| 32. Clarity of minor themes | Minor themes are also presented in the findings. |
