## Appendix Table 2 for "Coproducing a conceptual understanding of unmet palliative care needs: stakeholder workshops using modified nominal group technique"

**Appendix Table 2 Elements of unmet palliative care needs, generated inductively from stakeholder workshops with people with lived experience and people with professional experience, prior to further refinement from stakeholders**

| | |
|---|---|
| A | Unaddressed pain |
| B | Unaddressed breathlessness |
| C | Other unaddressed physical symptoms |
| D | Lacking support for functional independence |
| E | Unaddressed depression |
| F | Unaddressed anxiety |
| G | Feeling unsafe and unsupported |
| H | Unaddressed spiritual needs |
| I | Unaddressed cultural needs |
| J | Unaddressed housing, financial or other practical issues |
| K | Not being treated like a person |
| L | Lack of timely assessment of what is important to the person |
| M | Lack of timely access to medicines |
| N | Lack of single point of contact to access support including out of hours |
| O | Lack of coordination and continuity of care |
| P | Inability to access services needed |
| Q | Lack of referral or access to specialist palliative care if needed |
| R | Lack of information about disease and treatment choices (including self-management) |
| S | Lack of information about services and support available |
| T | Lack of access to equipment (e.g. walking aids, commodes) |
| U | Lack of help with personal care |
